# Human metapneumovirus-associated hospitalisation burden in children aged under 5 years in Scotland between 2017 and 2023: a retrospective analysis

**DOI:** 10.64898/2026.03.22.26349006

**Authors:** Durga Kulkarni, Richard Osei-Yeboah, Kate Templeton, Harish Nair

## Abstract

**Background:** Human metapneumovirus (hMPV) is commonly associated with respiratory tract infections (RTIs) in young children.

**Methods:** We estimated the annual hospital incidence of hMPV RTIs in children under 5 years in Scotland from 2017 to 2023 using national hospital and laboratory data. Incidence outside Lothian, where testing practices were uncertain, was extrapolated from Lothian laboratory data, where hMPV testing was advised for all RTI admissions. We also examined the severity and mortality of laboratory-confirmed hMPV cases. We developed similar estimates for RSV and Influenza A for comparison.

**Results:** This analysis included 1,462 laboratory-confirmed hMPV hospitalisations in children aged under 5 years. The extrapolated hMPV hospital incidence ranged from 19 per 100,000 to 537 per 100,000 in children aged under 5 years. The extrapolated incidence was two to three times higher than that based on laboratory-confirmed data. Hospital incidence was higher in infants than in toddlers. hMPV incidence dropped substantially during the 2020/21 season, followed by a rebound during the 2021/22 season. About 10% of hMPV RTI hospital admissions required hospital stay ≥5 days, but <1% required intensive care unit admissions or resulted in in-hospital death. RSV hospital incidence appeared substantially higher than the hMPV hospital incidence in this population.

**Conclusions:** hMPV RTIs contribute to a substantial hospital burden in young children in Scotland. However, the RSV RTI burden is likely to be higher in the population unvaccinated against both viruses. Improved surveillance and diagnosis strategies are required to develop robust hospital burden estimates.

**Summary:** Human metapneumovirus was associated with substantial hospitalisations in children under 5 years, particularly infants, in Scotland between 2017 and 2023. Incidence peaked in 2021/22 after a sharp decline in 2020/21. Most admissions were brief, with few requiring intensive care or resulting in death.

## Introduction

Human metapneumovirus (hMPV) is a respiratory paramyxovirus disproportionately affecting young children, older adults, and immunocompromised individuals (1). In young children, hMPV commonly presents as bronchiolitis and pneumonia (2). In 2018, hMPV was estimated to cause 14.2 million acute lower respiratory infection episodes worldwide (Uncertainty range [UR]: 10.2–20.1 million), leading to 643,000 hospitalisations (UR: 425,000–977,000) and 7,700 in-hospital deaths (UR: 2,600–48,800) (3).

Currently, licensed vaccines or targeted therapies for hMPV are unavailable. As a result, clinical management remains supportive, and routine testing is uncommon (4). This limits the ability to accurately estimate the national burden of hMPV-associated respiratory tract infections (RTIs). Quantifying this burden is, however, essential given the ongoing development of several hMPV vaccine candidates, which are undergoing testing in adults and children (5-7). The recent licensure of RSV vaccines in the UK further underscores the need to assess their potential impact on hMPV epidemiology in young children, as RSV and hMPV are closely related respiratory viruses with some overlapping epidemiological and clinical characteristics. Such assessments require an understanding of the baseline hospital burden of hMPV before the introduction of RSV vaccines.

Therefore, we estimated the hospital burden associated with hMPV-related RTIs among children younger than 5 years in Scotland across six annual seasons, from 1 July 2017 to 30 June 2023, using routinely collected hospital and laboratory data. By spanning the COVID-19 pandemic, this study also offers insights into shifts in hMPV epidemiology during this period. These findings contribute to monitoring disease trends and can help inform future hMPV immunisation policy in the United Kingdom.

## Methods

### Study design and setting

We performed a retrospective analysis of hMPV-associated RTI hospitalisations using data from Scottish national hospital registries for the period between July 1, 2017, and June 30, 2023, focusing on children under 5 years of age in Scotland. We applied a similar analytical approach as in our previous work examining hMPV burden in older adults in Scotland, with stratification and outcomes tailored to the under-5 years population (8).

The 2023 mid-year population of Scotland (across all ages) was approximately 5.4 million, with about 250,000 in the <5 years age band (9). The Lothian Health Board constitutes one of the fourteen NHS health boards in Scotland. It serves the second-largest residential population in the country, covering the councils of the City of Edinburgh, East Lothian, Midlothian, and West Lothian. The 2023 mid-year population of Lothian (across all ages) was approximately 0.9 million, with about 42,000 in the <5 years age band (9, 10).

Compared with other Scottish health boards, Lothian has a more ethnically diverse profile (Supplementary Table S1) (11). It also has a lower share of its population residing in the 10% most deprived areas, indicating lower overall deprivation levels (12).

Scotland has a universal, publicly funded healthcare system, and hospital care is free at the point of use for all residents. There were no vaccines against hMPV or RSV implemented through the national immunisation programme during the study period.

### Case definitions

We defined RTI hospitalisations as any hospital admission episode that included at least one RTI-related diagnostic code, recorded as either the primary or a secondary diagnosis, according to the International Classification of Diseases, 10^th^ edition (ICD-10) (13). Up to six diagnosis codes were available for each episode. The list of RTI codes is attached in Supplementary Table S2. We included all RTI hospitalisations noted as inpatient admissions, also including those with a duration of less than one day. Admissions recorded as day case admissions (admissions to a specialty for clinical care that involve clinical intervention and a period of supervised recovery within the treatment area) were excluded. Such day case patients are not expected to stay in the hospital overnight. Routine admissions were also excluded, as these represent planned hospital stays that are unlikely to be associated with the RTI episodes.

hMPV-specific RTI episodes were defined as RTI hospital episodes with a positive hMPV test on a specimen collected within 7 days and up to 3 days after the date of hospital admission. We included this window of 7-day pre-admission and 3 days post-admission to include test results before admission relevant to the hospital admissions based on the known incubation period for hMPV (3 to 6 days) and to include all relevant hospitalisation episodes, even when testing did not occur at the point of admission but subsequently during hospitalisation, while ruling out nosocomial infections, respectively.

The health board indicated which NHS health board the hospital, where the admission occurred, was part of.

We defined a season as the period between 1 July of one year and 30 June of the subsequent year, to capture all hospitalisations during the winter period.

Hospital length of stay (LOS) was defined as the number of days from admission to discharge. A long LOS was defined as a LOS ≥ the 90th percentile for hMPV-related RTIs in children under 5 years, corresponding to the top 10% of admissions during the study period.

Intensive care unit (ICU) admissions included ICU as well as high-dependency unit (HDU) admissions occurring within the hMPV RTI hospital admission episodes. ICU admission rate referred to the proportion of hMPV RTI admissions requiring ICU (or HDU) admission and was expressed as a percentage.

The in-hospital case fatality rate (CFR) was defined as the proportion of in-hospital deaths among all hMPV RTI admissions. Similarly, the post-discharge CFR within 90 days was defined as the proportion of deaths occurring after discharge and up to 90 days post-discharge among all hMPV RTI admissions. The CFRs were also expressed as percentages.

### Data sources and data linking

Hospitalisation episode data were extracted from the Scottish Morbidity Records 01 (SMR01). SMR01 is an episode-based patient record related to all in-patients and day case episodes from non-obstetric and non-psychiatric specialties in Scotland (14).

hMPV laboratory data were obtained from the Electronic Communication of Surveillance Scotland (ECOSS), a Public Health Scotland (PHS) system that collects laboratory test results, including patient identifiable data, from all NHS laboratories in Scotland (15). We could access only positive results without data on total or negative tests. These tests were part of routine standard of care. All RTI hospital episodes in Lothian during the study period were advised to undergo respiratory virus testing using the LUMINEX Respiratory Virus Panel, which included hMPV, RSV and Influenza A. In other health boards, testing policies were inconsistent and unclear, and varied by virus.

ICU or HDU admission data were sourced from the Scottish Intensive Care Society Audit Group (SICSAG), and mortality data were sourced from the National Records of Scotland (NRS) (16, 17).

We matched all datasets using a pseudonymised patient-identifiable number. Furthermore, the hospital admission (SMR01), laboratory (ECOSS) and ICU (SICSAG) data were also linked according to the hospital admission date, specimen collection date (as mentioned above) and ICU admission dates (ICU admission within the hospital stay).

Population estimates required for estimating population-based rates were obtained from the National Records of Scotland (9, 18). For each season, the mid-year population estimates at the end of the season were used.

## Ethical approval

The Public Benefit and Privacy Panel for Health and Social Care (PBPP-HSC) approved data access to the required data in the National Safe Haven (Approval number: 2223-0019).

### Patient consent

This research only examined existing data that were pseudonymised before analysis, and it did not require the active consent of patients. The data were collected routinely through NHS Scotland, with patients informed of potential use and their rights through various PHS and NHS Scotland privacy notices.

### Statistical analysis

We estimated the annual laboratory-confirmed hMPV-associated RTI hospital incidence for children under 5 years in Scotland over the six seasons (Supplementary Figure S1a). We calculated the 95% CI using non-parametric bootstrap resampling. We also developed similar estimates by stratifying the data into three age bands of 0-5 months, 6 months-1 year and 2-4 years.

Additionally, we extrapolated for under-testing of RTI hospital admissions in health boards other than Lothian using Lothian as a benchmark. For each month during the study period, we calculated the hMPV proportion positive among all the RTI-associated hospital admissions in Lothian, stratified by age bands (0-5 months, 6 months-1 year, and 2-4 years). These hMPV proportions positive among all-cause RTI estimates from Lothian were applied to the corresponding RTI admissions in other health boards. Finally, we summed up the corresponding estimates for each age band and month from Lothian and other health boards, during each season, to develop the extrapolated hospitalisation incidence estimates for the annual season (Supplementary Figure S1b). Again, the 95% CI was estimated using non-parametric bootstrap resampling.

We also conducted a sensitivity analysis and estimated the laboratory-confirmed hMPV hospital incidence in Lothian, limiting the inclusion to episodes where a specimen was collected for testing within 3 days before and 3 days after the admission date to exclude cases attributable to sequelae or complications.

We estimated the proportion of episodes requiring long LOS, by age bands, in Lothian and other health boards. The ICU admission rate, in-hospital CFR, and post-discharge CFR were expressed as percentages with 95% CI (bootstrap resampling method) for those under 5 years in Scotland. Stratification by age bands and regions was not possible due to low (<5) counts in several groups, leading to unstable estimates and data protection concerns.

We developed similar estimates for RSV and Influenza A to provide initial insights into trends across the three viruses in this age group.

## Results

Our analysis included 1,462 unique hospitalisation episodes associated with laboratory-confirmed hMPV RTIs in children aged under 5 years in Scotland between July 1, 2017, and June 30, 2023. The demographic characteristics of all-cause RTI hospital admissions and laboratory-confirmed hMPV RTI hospital admissions in <5 years in Scotland during the study period are provided in Table 1. Characteristics stratified by Lothian and other health boards are shown in Supplementary Table S3. Only 357 (24%) of the hospital episodes had hMPV-specific diagnosis codes (J12.3 or J21.1), either as primary or secondary diagnoses.

**Table 1:**
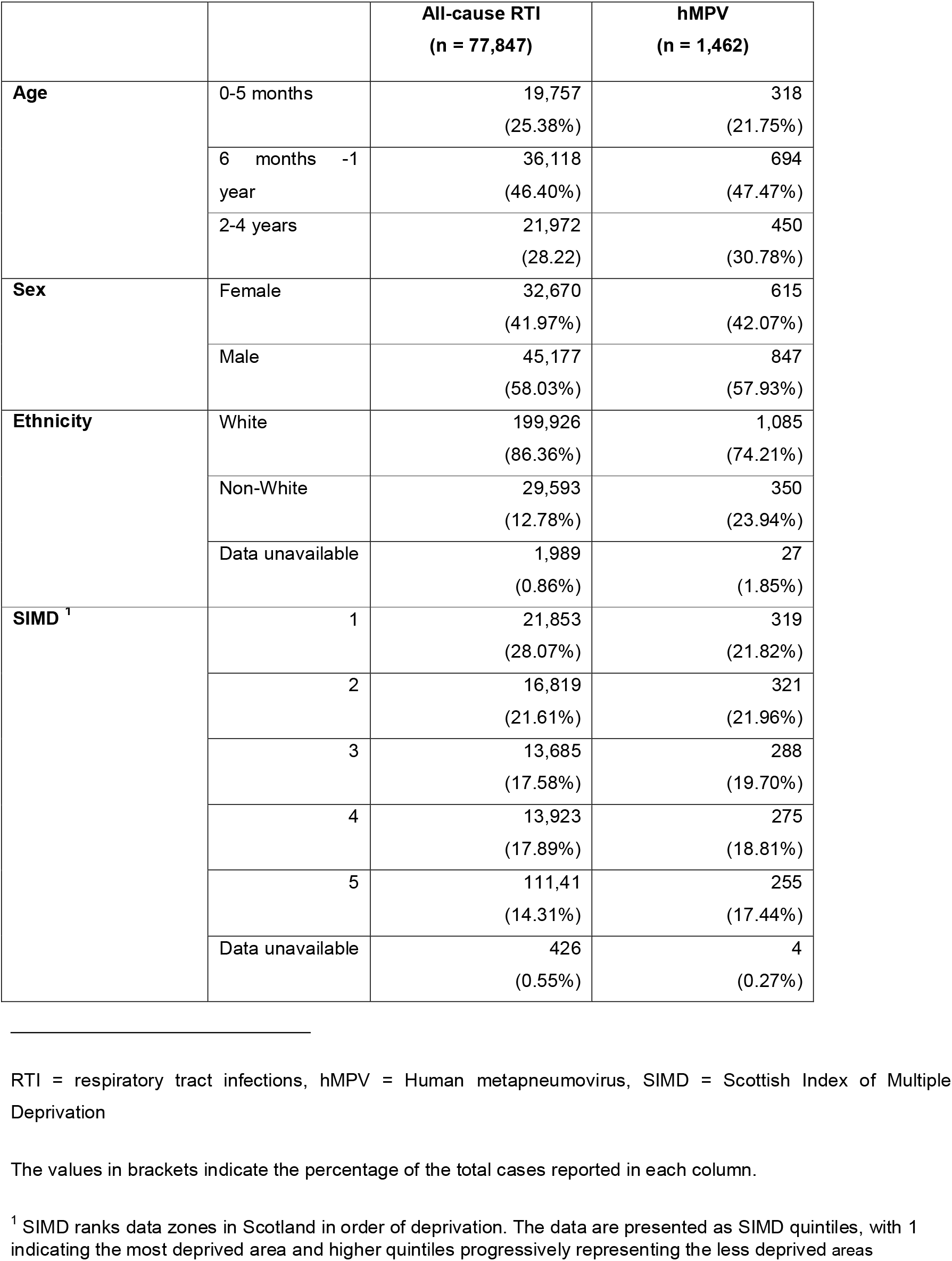
Characteristics of the study population.

Table 2 shows the annual laboratory-confirmed and extrapolated hospital incidence of hMPV RTIs in children under 5 years in Scotland, stratified by age groups (0–5 months, 6 months– 1 year, and 2–4 years) during the study period. We estimated that there were 756; 1,751; and 1,261 hospitalisation episodes in the 0–5 months, 6 months–1 year, and 2–4 years age bands, respectively, with these extrapolated estimates approximately 2.4, 2.5, and 2.8 times higher than the corresponding laboratory-confirmed episodes. Incidence could not be extrapolated for the 0–5 months age band in the 2020/21 season due to the absence of laboratory-confirmed cases in Lothian during that season. Laboratory-confirmed incidence in children under 5 years ranged from 9.2 (95% Confidence Interval [CI]: 6.0–13.1) to 225.7 (205.9–244.2) per 100,000 persons, while extrapolated incidence ranged from 19.2 (13.9– 24.7) to 537.0 (507.0–566.0) per 100,000 persons. The highest incidence occurred in the 2021/22 season, and the lowest in 2020/21 (both laboratory-confirmed and extrapolated estimates). Thus, the lowest incidence was observed during the first winter of the COVID-19 pandemic, while the highest incidence occurred in the subsequent season.

**Table 2:** Laboratory-confirmed and extrapolated annual hospital incidence of hMPV-associated RTIs (per 100,000) with 95% CI in children aged < 5 years in Scotland by age bands during six annual seasons (2017-2023)

| Age band | 0-5 months |  | 6 months – 1 year |  | 2-4 years |  | <5 years |  |
| --- | --- | --- | --- | --- | --- | --- | --- | --- |
| Season | Lab-confirmed annual hospital incidence | Extrapolated annual hospital incidence | Lab-confirmed annual hospital incidence | Extrapolated annual hospital incidence in 6m-1y | Lab-confirmed annual hospital incidence | Extrapolated annual hospital incidence | Lab-confirmed annual hospital incidence | Extrapolated annual hospital incidence |
| 2017/18 | 108.6<br>(81.4 – 135.7) | 180.4<br>(143.5 – 217.2) | 231.0<br>(189.4 – 272.7) | 646.6<br>(579.5 – 719.6) | 41.3 (31.7 – 52.0) | 129.9<br>(114.2 – 147.7) | 90.9 (79.9 – 102.3) | 240 (221.6 – 258.8) |
| 2018/19 | 150 (116.4 – 183.6) | 396.5<br>(341.4 – 454.0) | 256.6<br>(214.2 – 303.0) | 790.2<br>(717.8 – 874.1) | 48.0 (37.7 – 59.0) | 168.0<br>(148.3 – 188.5) | 107.9<br>(95.1 – 120.2) | 332.2<br>(312.0 – 354.7) |
| 2019/20 | 99.1 (68.1 – 128.0) | 246.6<br>(218.8 – 313.8) | 192.2<br>(154.9 – 231.4) | 432.3<br>(378.5 – 488.3) | 36.9 (28.1 – 45.7) | 97.1 (81.3 – 112.6) | 79.0 (68.2 – 89.1) | 194.3<br>(178.5 – 211.3) |
| 2020/21 | 8.6 (2.1 – 19.3) | * | 22.6 (10.3 – 37.0) | 56.5 (37.0 – 76.2) | 5.1 (1.9 – 9.0) | 10.7 (5.8 – 16.1) | 9.2 (6.0 – 13.1) | 19.2 (13.9 – 24.7) |
| 2021/22 | 254.3<br>(209.8 – 298.8) | 666.9<br>(595.5 – 741.7) | 539.6<br>(474.5 – 606.8) | 1201.4<br>(1102.3 – 1299.6) | 119.0<br>(101.4 – 136.7) | 290.0<br>(262.2 – 315.9) | 225.7<br>(205.9 – 244.2) | 537.0<br>(507.0 – 566.0) |
| 2022/23 | 30.3 (17.3 – 47.6) | 31.4 (17.3 – 47.6) | 150.6<br>(115.5 – 183.6) | 374.1<br>(321.8 – 431.2) | 34.7 (25.6 – 44.6) | 100.5<br>(85.2 – 116.6) | 56.6 (47.7 – 66.3) | 141.2<br>(127.8 – 155.7) |

Across all seasons, incidence was highest in children aged 6 months–1 year. The second-highest incidence was observed in infants aged 0–5 months during the 2017/18–2021/22 seasons (first five seasons), and in children aged 2–4 years in the 2022/23 season (Table 2).

The all-cause RTI hospital incidence and the monthly laboratory-confirmed proportion positives in RTI in Lothian, both of which were used in the extrapolation, are provided in Supplementary Tables S4 and S5, respectively. We have also provided the laboratory-confirmed hospital incidence and the extrapolated hospital incidence stratified by Lothian and other health boards in Supplementary Table S6.

The sensitivity analyses conducted by limiting episodes to specimens collected within 3 days before and 3 days after the date of admission in Lothian demonstrated that the estimates were similar to the main analysis (with the 7-day cut-off pre-admission). The results of this sensitivity analysis are reported in Supplementary Table S7.

Table 3 shows the proportion of hospital episodes with a prolonged LOS (≥5 days) by age group in Lothian and in other Scottish health boards. Overall, a higher proportion of hMPV-related RTI episodes required long LOS in Lothian compared with other health boards among children under 5 years. The proportion requiring long LOS was comparable across the three age bands in Lothian but more variable in other health boards.

**Table 3:** Number and proportion of laboratory-confirmed hMPV RTI episodes requiring long hospital length of stay in children aged < 5 years in Scotland by age bands during six annual seasons (2017-2023)

| Age bands | Lothian | Other health boards | Overall (Scotland) |
| --- | --- | --- | --- |
| **0-5 months** | 16 (13.1%) | 20 (10.2%) | 36 (11.3%) |
| **6 months – 1 year** | 35 (15.2%) | 35 (7.5%) | 70 (10.1%) |
| **2-4 years** | 19 (15.0%) | 19 (6.4%) | 38 (8.4%) |
| **Overall (<5 years)** | 70 (13.9%) | 74 (7.7%) | 98 (9.8%) |

The ICU admission rate was 0.34% (95% CI: 0.07 – 0.7) in children aged under 5 years in Scotland between 2017 and 2023. The in-hospital CFR was 0.14% (95% CI: 0.00 – 0.34) and the post-discharge CFR within 90 days was 0.34% (95% CI: 0.07 – 0.7) in this population and during this period.

There were 10,985 laboratory-confirmed RSV and 2,124 laboratory-confirmed Influenza A associated unique hospital admissions in children aged under 5 years during the study period in Scotland. The demographic characteristics of laboratory-confirmed RSV and Influenza A episodes are presented in Supplementary Table S8. About 65% and 58%, respectively, of these RSV and Influenza A episodes had virus-specific diagnostic codes, either as primary or secondary diagnoses. Across the three age bands, both laboratory-confirmed and extrapolated hospital incidence appeared to be highest for RSV, with some inconsistent patterns between Influenza A and hMPV. About 8% of RSV and 5% of Influenza A hospital admissions required a LOS ≥5 days (Supplementary Table S9). ICU admission rates and CFR estimates for the three viruses had overlapping 95% CIs. However, these estimates were based on low counts (often <5) and, therefore, subject to considerable uncertainty (Supplementary Table S10).

## Discussion

Our study showed that hMPV-associated RTI hospitalisations in children aged under 5 years, especially in infants aged less than 1 year, impose a substantial hospital burden in Scotland. The COVID-19 pandemic coincided with atypical epidemiological trends, including an unusually low-incidence season followed by a rebound period and a shift in the age distribution. About 10% of hMPV RTI hospital episodes required a length of stay longer than 5 days. The ICU admission rate was 0.34%, and the in-hospital CFR was 0.14% in children aged under 5 years in Scotland between 2017 and 2023.

Our findings indicate that the highest burden of hMPV-related RTI hospitalisations occurred in children aged 6 months–1 year (Table 2). Several factors may explain the increased burden observed in late infancy during the pre-COVID-19 pandemic period. First, the combined effects of immunological immaturity and the progressive decline of maternally derived antibodies likely increase susceptibility to hMPV-associated RTIs in this age group. Evidence suggests that seroprevalence of hMPV-specific antibodies exceeds 90% in infants younger than 3 months, indicating substantial passive protection early in life (19). By 6–12 months of age, maternal antibody levels have largely waned, while infection-induced immunity remains limited, creating a period of increased vulnerability to clinically significant hMPV infections. Second, increased opportunities for viral exposure in later infancy, resulting from expanded social contact networks compared with the more protected neonatal period, may further contribute to this pattern. Third, given the established clinical importance of RSV in early infancy and the incomplete testing strategies in health boards other than Lothian, hMPV infections in infants aged 0–5 months may have been under-detected if diagnostic efforts were preferentially directed towards RSV.

We attempted to account for undertesting by developing extrapolated hospital incidence estimates. Even after extrapolation, the 6 months–1 year age band continued to have higher incidence estimates than the 0–5 months and 2–4 years groups, indicating that differential undertesting across the two youngest age bands is unlikely to fully explain the observed age trend (Table 2). The pattern of highest hospital incidence in infants aged 6 months–1 year, followed by those aged 0–5 months in the five seasons, is broadly consistent with published evidence that hMPV predominantly affects infants, who experience substantially higher hospitalisation burden than older children (3). However, our previous systematic review at global level, based on published data from high-income countries, indicated higher overall admission rates in younger infants aged 0–5 months (330 per 100,000) compared with those aged 6 months–1 year (280 per 100,000), whereas studies from low- and middle-income countries tend to report higher rates in the 6 months–1 year age band (270 per 100,000) than in infants 0–5 months (240 per 100,000) (3). These comparisons suggest that while the broader trend of higher burden in infants is consistent, differences observed within narrower infant age bands in our study may reflect population-level variations. Moreover, the systematic review showed overlapping 95% confidence intervals for the two age bands, likely suggesting insignificant differences, and the current study findings may have been influenced by incomplete testing.

Our study demonstrated a sharp decline in hMPV-related hospitalisations during the 2020/21 season (Table 2). While extrapolated incidence among children under 5 years ranged between 190 and 240 per 100,000 persons in the preceding three seasons, it fell to 9.4 per 100,000 in 2020/21. In the 2021/22 season, when COVID-19 non-pharmacological interventions (NPIs) were gradually relaxed, hospital incidence rebounded to 537 per 100,000 persons in the under-5 population. This rebound likely reflects the ‘immunity debt’ accumulated during the pandemic, resulting in an unprecedented rise in symptomatic infections from non-SARS-CoV-2 respiratory viruses when NPIs were eased (20). Interestingly, there was also a shift in the age distribution during the 2022/23 season. Although children aged 6 months–1 year continued to have the highest hospital incidence, those aged 2–4 years surpassed infants aged 0–5 months. This atypical shift likely reflects post-pandemic changes in population immunity and delayed viral exposure among young children due to COVID-19 non-pharmaceutical interventions, which are expected to be temporary. This unusual shift likely reflects transient post-pandemic changes in population immunity and delayed viral exposure among young children resulting from COVID-19 NPIs.

Long LOS (≥5 days) was observed more frequently in Lothian, despite lower overall socio-economic deprivation and a generally healthier population demographic (Table 3, Supplementary Table S1). This may not necessarily indicate greater disease severity in Lothian. In addition to severity, LOS can be influenced by admission and discharge practices, inpatient resource availability, local care pathways, and healthcare accessibility. Higher hospital density in Lothian, implying greater bed capacity, may be associated with comparatively lower bed pressures and more conservative discharge practices, particularly for infants, potentially contributing to longer lengths of stay despite an overall healthier population (21).

Although ICU admission rates and in-hospital case-fatality appeared low, the hospital incidence indicates a substantial burden of symptomatic disease requiring hospital care. hMPV-associated admissions can strain hospital bed capacity, especially during peak respiratory seasons. We were unable to estimate ICU admission rates and CFRs stratified by narrower age bands or health boards (Lothian vs others) due to low counts (<5) in several groups. Consequently, incomplete testing policies outside Lothian may have biased these estimates. While selective testing of severe cases may have overestimated ICU admission rates or CFRs, incomplete testing could have led to under-ascertainment of laboratory-confirmed hMPV infections and severe cases. Therefore, predicting the direction or magnitude of the resulting bias is challenging.

Across the six consecutive seasons between 1 July 2017 and 30 June 2023, the extrapolated hospital incidence of RSV appeared to be consistently highest in the under-5 years, except in the 2020/21 season, when hMPV seemed to surpass RSV (Supplementary Table S4). During the pre-pandemic seasons (2017/18 to 2019/20), hMPV hospital incidence estimates were 4 to 8-fold lower than RSV hospital incidence estimates in this population. The unusual shift (RSV vs hMPV) during the 2020/21 season coincided with an unusual age trend and a high incidence of hMPV during the COVID-19 pandemic, as described earlier. While RSV generally had the highest extrapolated hospital incidence in children under five, the relative ranking of influenza A and hMPV varied by season and age group (Supplementary Table S4). Incomplete and differential virus-specific coding, particularly the under-coding of hMPV, limits the reliability of ICD-based comparisons and has important implications for surveillance, research using clinically coded hospital data, and future burden-of-disease estimates. A seemingly higher proportion of hMPV RTI hospital admissions had a length of stay ≥5 days compared with RSV and Influenza A; however, differences were not tested for statistical significance due to variable testing practices across viruses (Supplementary Table S9). Similarly, ICU admission rates and case fatality outcomes could not be formally compared because of inconsistent testing and low event counts. These findings highlight the need for further research using robust, comparable data to evaluate disease severity and outcomes across respiratory viruses.

We acknowledge that our study has a few limitations. Not all RTI hospitalisations underwent laboratory testing for hMPV across Scotland, so the incidence of laboratory-confirmed hospitalisations underestimates the true burden. Residual biases affecting hMPV RTI hospitalisation incidence, even after extrapolation, can be grouped into two categories: biases in RTI ascertainment and biases in hMPV ascertainment. For RTI-related biases, demographic differences between Lothian and other health boards may have affected all-cause RTI data. Our extrapolation accounted for age by stratifying the <5 years population into three narrower age bands, but we could not adjust for ethnicity, household composition or SIMD, as sample sizes were too small after stratification. The complex interactions between incidence, healthcare access, and health behaviours make it difficult to predict the overall impact of these factors on RTI ascertainment, potentially resulting in residual biases. Biases specific to hMPV include potential differences in infection risk within RTI populations and fluctuations caused by local outbreaks. Extrapolating from Lothian to other health boards may have over- or underestimated the incidence depending on outbreaks associated with hMPV or other viruses in Lothian or any of the other health boards. Finally, we aggregated monthly rather than weekly hMPV-positive RTI proportions due to low weekly case counts. This may have obscured short-term fluctuations in incidence, causing under- or overestimation of short-term variation.

## Conclusions

hMPV-associated RTIs represent a substantial hospital burden among children under 5 years in Scotland. Incomplete testing limited formal statistical comparisons with RSV, nevertheless, our findings indicate a notable age-specific burden that can help guide resource allocation and paediatric respiratory service planning. The COVID-19 pandemic was associated with atypical epidemiological patterns, including a low-incidence period, a subsequent rebound, and shifts in age distribution. Improved longitudinal surveillance with broader testing is needed to enable subgroup analyses, assess the impact of age and other demographic factors on disease severity, and inform the evaluation and targeting of future hMPV vaccination strategies.

## Data availability statement

The individual-level data underlying this article cannot be shared publicly, as they were provided by Public Health Scotland under approval from the Public Benefit and Privacy Panel for Health and Social Care and accessed via a secure platform.

## Acknowledgements

The authors would like to acknowledge the support of the eDRIS Team (Public Health Scotland) for their involvement in obtaining approvals, provisioning and linking data and the use of the secure analytical platform within the National Safe Haven.

## Author contributions

DK, HN, and KT conceived the study idea. HN supervised the study. DK led and conducted the data cleaning, data linking and data analysis with significant contributions from ROY and HN. DK wrote the first draft of the manuscript. All authors provided critical revisions to the manuscript and approved the final version.

