## Supplementary material for "Human metapneumovirus-associated hospitalisation burden in children aged under 5 years in Scotland between 2017 and 2023: a retrospective analysis"

### Supplementary Table S1: Underlying population characteristics in Lothian and other Scottish health boards in 2022 * †

| | Lothian | Other Scottish health boards | Scotland |
|---|---|---|---|
| **Total population (all ages)** | 904,628 | 4,535,214 | 5,439,842 |
| **0-5m** | 8,390 (0.93%) | 39,354 (0.87%) | 47,744 (0.88%) |
| **6m-1y** | 8,198 (0.91%) | 38,501 (0.85%) | 46,699 (0.86%) |
| **2-4y** | 25,626 (2.83%) | 128,336 (2.83%) | 153,962 (2.83%) |
| **Overall <5 years** | 42,214 (4.66%) | 206,191 (4.55%) | 248,405 (4.57%) |
| **White ethnicity** | 809,550 (89.49%) | 4,242,323 (93.5%) | 5,051,873 (92.87%) |
| **Percentage of health board population in the 10% most deprived areas (SIMD 2020)** | 4.2% | -- | 10% |

*National Records of Scotland. Ethnic group, national identity, language and religion 2025 [Available from: https://www.scotlandscensus.gov.uk/search-the-census#/topics/list?topic=Ethnic%20group,%20national%20identity,%20language%20and%20religion&categoryId=1

† Public Health Scotland. Deprivation: data 2025 [Available from: https://www.scotpho.org.uk/wider-determinants/deprivation/data/.

Lothian ranks 8th among 14 Scottish Health Boards for the proportion of its population in the 10% most deprived areas

### Supplementary Table S2: ICD-10 respiratory tract infection diagnostic codes

| Condition | ICD-10 codes |
|---|---|
| Acute respiratory tract infections | J00\|J02\|J020\|J028\|J029\|J03\|J038\|J039\|J04\|J040\|J041\|J042\|J05\|J050\|J051\|J06\|J060\|J068\|J069\|J00\|J02\|J02.0\|J02.8\|J02.9\|J03\|J03.8\|J03.9\|J04\|J04.0\|J04.1\|J04.2\|J05\|J05.0\|J05.1\|J06\|J06.0\|J06.8\|J06.9 |
| Pneumonia and Influenza codes | J110\|J111\|J12\|J120\|J121\|J12\|J128\|J129\|J178\|J18\|J180\|J181\|J182\|J188\|J189\|J11.0\|J11.1\|J12\|J12.0\|J12.1\|J12\|J12.8\|J12.9\|J17.8\|J18\|J18.0\|J18.1\|J18.2\|J18.8\|J18.9 |
| Bronchiolitis and bronchitis | J20\|J200\|J201\|J202\|J206\|J207\|J208\|J209\|J21\|J218\|J219\|J40\|J20\|J20.0\|J20.1\|J20.2\|J20.6\|J20.7\|J20.8\|J20.9\|J21\|J21.8\|J21.9\|J40 |
| Unspecified LRTI | J22 |
| Human metapneumovirus codes | J123\|J211\|J12.3\|J21.1 |
| Respiratory syncytial virus codes | J121\|J205\|J210\|B974\|J12.1\|J20.5\|J21.0\|B97.4 |
| Flu codes | J09\|J10\|J100\|J101\|J108\|J11\|J11.8\|J09\|J10\|J10.0\|J10.1\|J10.8\|J11\|J11.8 |
| SARS-CoV-2 | U071\|U072\|U089\|U099\|U109\|U07.1\|U07.2\|U08.9\|U09.9\|U10.9 |

#### Supplementary Figure S1: Flow diagram describing the methods of estimating the laboratory-confirmed hospital incidence and extrapolated hospital incidence in Scotland

##### Supplementary Figure S1a: Hospital incidence for Scotland based on laboratory-confirmed data from all Scottish health boards (without extrapolation)

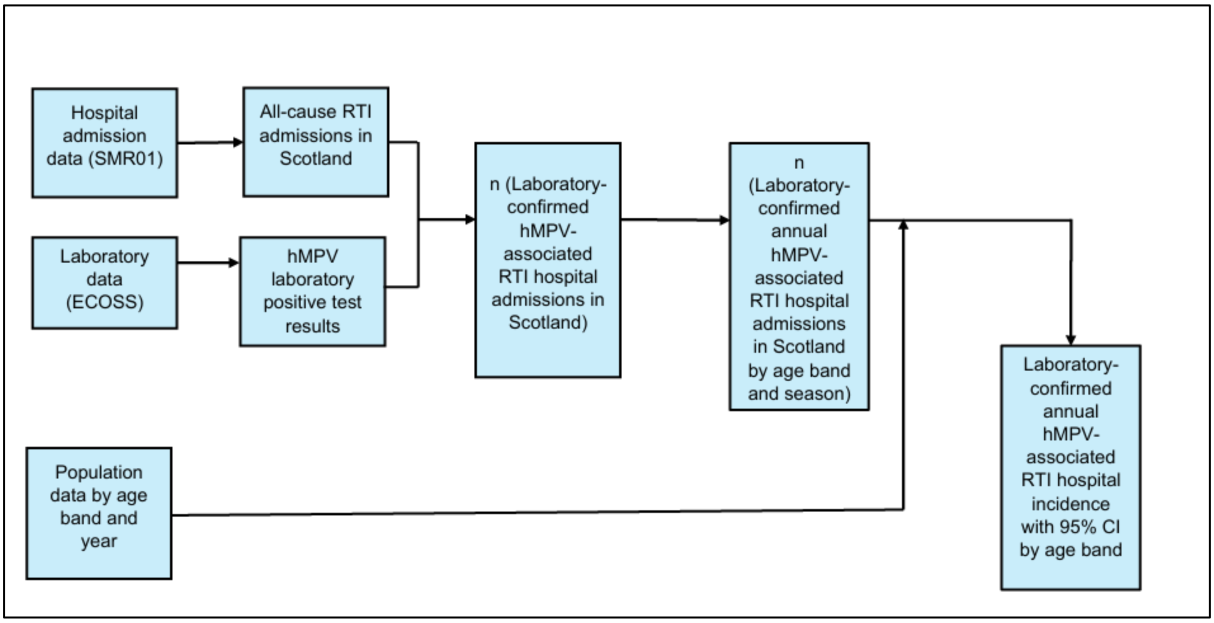

Supplementary Figure S1b: Hospital incidence for Scotland based on laboratory-confirmed data from Lothian and RTI hospital admissions in Scotland with extrapolation to health boards except Lothian

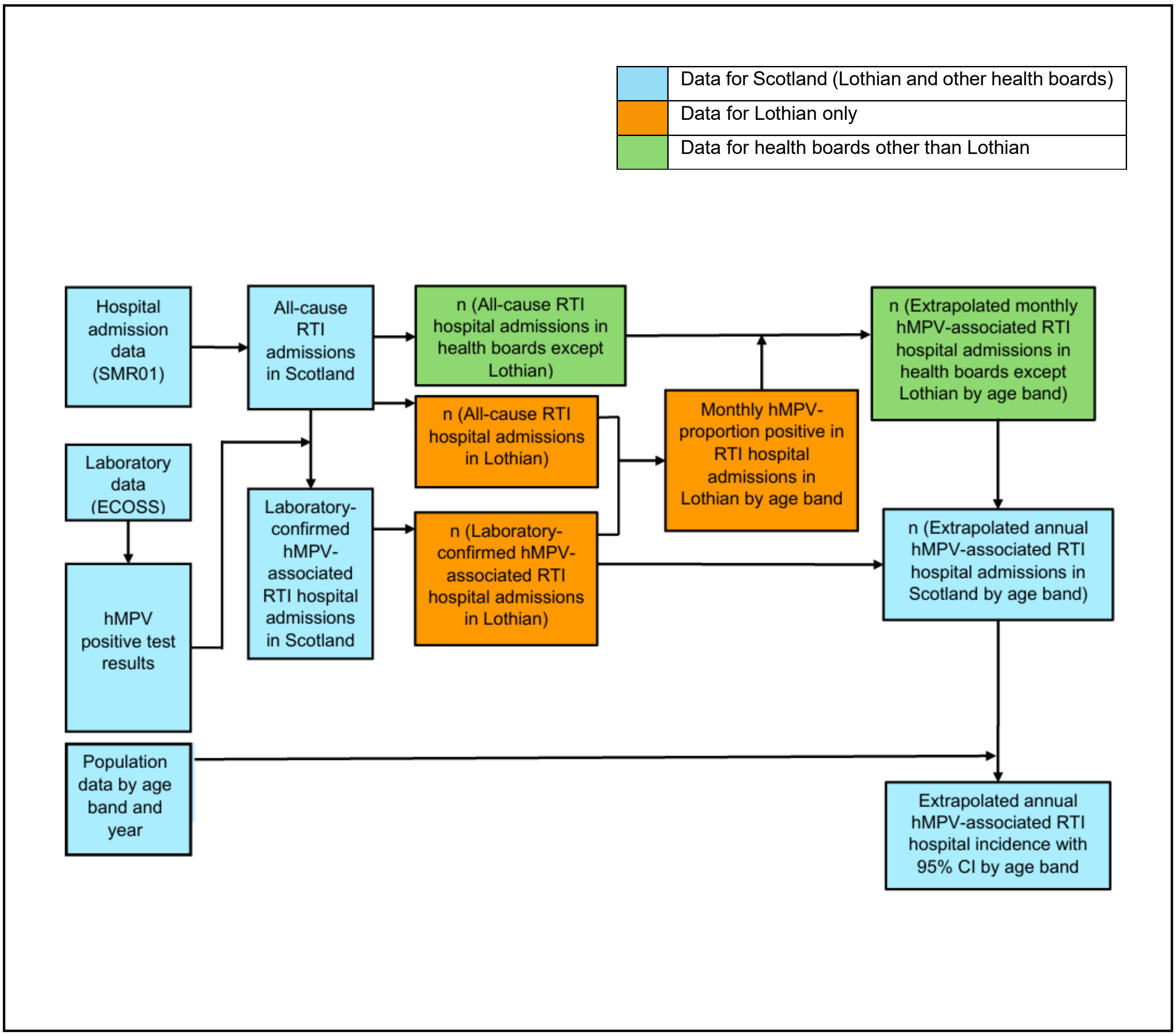

### Supplementary Table S3: Study population characteristics by health boards – Lothian and other Scottish health boards

| | | All-cause RTI in Lothian (n = 10,572) | All-cause RTI in other health boards (n = 67,275) | Laboratory-confirmed hMPV in Lothian (n = 505) | Laboratory-confirmed hMPV in other health boards (n = 957) | Laboratory-confirmed RSV in Lothian (n = 2,406) | Laboratory-confirmed RSV in other health boards (n = 8,579) | Laboratory-confirmed Influenza A in Lothian (n = 377) | Laboratory-confirmed Influenza A in other health boards (n = 1,747) |
|---|---|---|---|---|---|---|---|---|---|
| **Age** * | 0-5 months | 3,242 (30.7%) | 16,515 (24.5%) | 122 (24.2%) | 196 (20.5%) | 1,101 (45.8%) | 3,924 (45.7%) | 68 (18%) | 253 (14.5%) |
| | 6 months – 1 year | 4,723 (44.7%) | 31,395 (46.7%) | 230 (45.5%) | 464 (48.5%) | 971 (40.4%) | 3,324 (38.7%) | 156 (41.4%) | 735 (42.1%) |
| | 2-4 years | 2607 (24.7%) | 193,65 (28.8%) | 153 (30.3%) | 297 (31%) | 334 (13.9%) | 1,331 (15.5%) | 153 (40.6%) | 759 (43.4%) |
| **Sex** | Female | 4,442 (42%) | 28,228 (42%) | 209 (41.4%) | 406 (42.4%) | 1,072 (44.6%) | 3,795 (44.2%) | 157 (41.6%) | 792 (45.3%) |
| | Male | 6,130 (58%) | 39,047 (58%) | 296 (58.6%) | 551 (57.6%) | 1,334 (55.4%) | 4,784 (55.8%) | 220 (58.4%) | 955 (54.7%) |
| **Ethnicity †** | White | 8,388 (79.3%) | 47,124 (70%) | 408 (80.8%) | 677 (70.7%) | 1,892 (78.6%) | 5,649 (65.8%) | 287 (76.1%) | 1237 (70.8%) |
| | Non-White | 2,184 (21.7%) | 16,286 (24.2%) | 97 (19.2%) | 253 (26.4%) | 514 (21.4%) | 2,106 (24.5%) | 90 (23.9%) | 381 (21.8%) |
| | Data unavailable | 0 | 3,865 (5.7%) | 0 | 27 (2.8%) | 0 | 824 (9.6%) | 0 | 129 (7.4%) |
| **SIMD ‡** | 1 | 1,937 (18.3%) | 19,916 (29.6%) | 100 (19.8%) | 219 (22.9%) | 438 (18.2%) | 2,498 (29.1%) | 85 (22.5%) | 536 (30.7%) |
| | 2 | 2,585 (24.5%) | 14,234 (21.2%) | 113 (22.4%) | 208 (21.7%) | 614 (25.5%) | 1,721 (20.1%) | 86 (22.8%) | 392 (22.4%) |
| | 3 | 1,669 (15.8%) | 12,016 (17.9%) | 86 (17.0%) | 202 (21.1%) | 372 (15.5%) | 1,614 (18.8%) | 53 (14.1%) | 311 (17.8%) |
| | 4 | 2,291 (21.7%) | 11,632 (17.3%) | 102 (20.2%) | 173 (18.1%) | 520 (21.6%) | 1,516 (17.7%) | 77 (20.4%) | 277 (15.9%) |
| | 5 | 2,065 (19.5%) | 9,076 (13.5%) | 104 (20.6%) | 151 (15.8%) | 457 (19%) | 1,184 (13.8%) | 75 (19.9%) | 221 (12.7%) |
| | Data unavailable | 25 (0.2%) | 401 (0.6%) | 0 | 4 (0.4%) | 5 (0.2%) | 46 (0.5%) | 1 (0.3%) | 10 (0.6%) |

* The Chi-square test showed a statistically significant distribution in the age distribution between all-cause RTIs in Lothian and other health boards ($\chi^2$=26.75, p<0.001)
† The Chi-square test showed a statistically significant distribution in the ethnicity distribution between all-cause RTIs in Lothian and other health boards ($\chi^2$=2053.51, p<0.001)
‡ The Chi-square test showed a statistically significant distribution in the SIMD distribution between all-cause RTIs in Lothian and other health boards ($\chi^2$=5317.39, p<0.001)

### Supplementary Table S4: Laboratory-confirmed and extrapolated hospitalisation incidence of hMPV, RSV and Influenza A per 100,000 persons (with 95% CI) by age bands and season in <5y in Scotland

| Season | Age band | All-cause RTI hospital incidence | Lab-confirmed hMPV hospital incidence | Extrapolated hMPV hospital incidence | Lab-confirmed RSV hospital incidence | Extrapolated RSV hospital incidence | Lab-confirmed Influenza A hospital incidence | Extrapolated Influenza A hospital incidence |
|---|---|---|---|---|---|---|---|---|
| 2017/18 | 0-5m | 7228.3 (7017.0 - 7461.0) | 108.6 (81.4 - 135.7) | 180.4 (143.5 - 217.2) | 1696.6 (1584.1 - 1809.1) | 2238.8 (2115.3 - 2367.5) | 164.8 (129.9 - 197.8) | 313.1 (265.6 - 364.5) |
| 2018/19 | 0-5m | 7762.6 (7537.5 - 7987.7) | 150.0 (116.4 - 183.6) | 396.5 (341.4 - 454.0) | 1734.9 (1626.3 - 1855.3) | 2802.8 (2664.5 - 2948.8) | 171.7 (134.2 - 211.2) | 211.1 (169.7 - 250.7) |
| 2019/20 | 0-5m | 7641.7 (7412.5 - 7883.2) | 99.1 (68.1 - 128) | 264.6 (218.8 - 313.8) | 2470.8 (2336.7 - 2613.3) | 3243.5 (3085.9 - 3397.7) | 177.5 (138.3 - 218.8) | 270.6 (227.1 - 320.0) |
| 2020/21 | 0-5m | 2122.1 (1993.6 - 2248.4) | 8.6 (2.1 - 19.3) | * | 2.1 (0 - 6.4) | * | 2.1 (0 - 6.4) | * |
| 2021/22 | 0-5m | 10247.7 (9997.6 - 10525.3) | 254.3 (209.8 - 298.8) | 666.9 (595.5 - 741.7) | 2316.1 (2180.5 - 2434.8) | 3425.9 (3261.2 - 3598.1) | 38.1 (21.2 - 55.1) | 47.5 (29.6 - 67.9) |
| 2022/23 | 0-5m | 5556.6 (5363.8 - 5764.6) | 30.3 (17.3 - 47.6) | 31.4 (17.3 - 47.6) | 2121.3 (1991.4 - 2262.1) | 1878.6 (1751.2 - 2011.0) | 95.2 (69.3 - 123.4) | 92.2 (64.9 - 121.2) |
| 2017/18 | 6m-1y | 13855.9 (13573.6 - 14157.0) | 231.0 (189.4 - 272.7) | 646.6 (579.5 - 719.6) | 1374.8 (1270.6 - 1473.3) | 2233.8 (2105.7 - 2367.1) | 321.9 (274.5 - 369.3) | 457.4 (397.7 - 515.1) |
| 2018/19 | 6m-1y | 13771.9 (13484.3 - 14078.7) | 256.6 (214.2 - 303.0) | 790.2 (717.8 - 874.1) | 1574.6 (1466.5 - 1678.9) | 3284.9 (3131.8 - 3450.3) | 553.8 (490.1 - 617.5) | 944.8 (866.4 - 1030.4) |

* There were no laboratory-confirmed episodes in Lothian in the age band and season combination. As a result, extrapolation was not possible.

| Season | Age band | All-cause RTI hospital incidence | Lab-confirmed hMPV hospital incidence | Extrapolated hMPV hospital incidence | Lab-confirmed RSV hospital incidence | Extrapolated RSV hospital incidence | Lab-confirmed Influenza A hospital incidence | Extrapolated Influenza A hospital incidence |
|---|---|---|---|---|---|---|---|---|
| 2019/20 | 6m-1y | 13102.6 (12800.5 - 13408.6) | 192.2 (154.9 - 231.4) | 432.3 (378.5 - 488.3) | 2153.4 (2033.7 - 2282.9) | 3791.0 (3622.3 - 3971.4) | 423.6 (368.7 - 482.4) | 558.9 (494.2 - 625.7) |
| 2020/21 | 6m-1y | 6460.7 (6240.6 - 6678.7) | 22.6 (10.3 - 37.0) | 56.5 (37.0 - 76.2) | 8.2 (2.1 - 18.5) | * | 2.1 (0 - 6.2) | * |
| 2021/22 | 6m-1y | 15181.9 (14879.5 - 15511.8) | 539.6 (474.5 - 606.8) | 1201.4 (1102.3 - 1299.6) | 1725.8 (1618.7 - 1837.2) | 3289.3 (3136.7 - 3445.5) | 79.8 (54.6 - 105.0) | 104.6 (75.6 - 136.5) |
| 2022/23 | 6m-1y | 9512.1 (9256.3 - 9774.2) | 150.6 (115.5 - 183.6) | 374.1 (321.8 - 431.2) | 1710.2 (1586.3 - 1821.6) | 1874.7 (1763.8 - 2001.1) | 369.3 (315.6 - 422.9) | 394.5 (342.4 - 453.8) |
| 2017/18 | 2-4y | 2331.6 (2259.3 - 2404.0) | 41.3 (31.7 - 52.0) | 129.9 (114.2 - 147.7) | 135.1 (117.8 - 153.1) | 199.2 (177.0 - 223.6) | 76.5 (64.0 - 90.3) | 106.7 (90.9 - 123.2) |
| 2018/19 | 2-4y | 2519.4 (2442.8 - 2593.0) | 48.0 (37.7 - 59.0) | 168.0 (148.3 - 188.5) | 150.8 (132.5 - 170.8) | 338.7 (313.1 - 366.6) | 187.9 (166.0 - 210.4) | 354.1 (325.3 - 382.4) |
| 2019/20 | 2-4y | 2353.2 (2278.2 - 2425.8) | 36.9 (28.1 - 45.7) | 97.1 (81.3 - 112.6) | 193.9 (173.9 - 215.8) | 270.1 (244.5 - 297.7) | 115.7 (98.2 - 132.0) | 176.7 (155.7 - 198.3) |
| 2020/21 | 2-4y | 1120.5 (1072.3 - 1175.1) | 5.1 (1.9 - 9.0) | 10.7 (5.8 - 16.1) | 2.6 (0.6 - 5.1) | 7.5 (3.2 - 12.2) | 2.6 (0.6 - 5.1) | * |
| 2021/22 | 2-4y | 3256.2 (3165.3 - 3343.3) | 119.0 (101.4 - 136.7) | 290.0 (262.2 - 315.9) | 329.6 (302.1 - 358.4) | 552.8 (514.0 - 594.5) | 40.5 (31.4 - 50.4) | 60.5 (48.4 - 73.9) |
| 2022/23 | 2-4y | 2255.6 (2179.6 - 2327.7) | 34.7 (25.6 - 44.6) | 100.5 (85.2 - 116.6) | 244.4 (220.8 - 268.7) | 319.1 (290.3 - 346.7) | 146.8 (127.1 - 166.5) | 170.4 (150.1 - 190.7) |
| 2017/18 | <5y | 5501.6 (5417.7 - 5588.2) | 90.9 (79.9 - 102.3) | 240.0 (221.6 - 258.8) | 672.6 (642.0 - 702.0) | 982.0 (943.9 - 1020.1) | 141.0 (127.0 - 155.7) | 214 (196.6 - 231.9) |
| 2018/19 | <5y | 5698.7 (5614.4 - 5786.4) | 107.9 (95.1 - 120.2) | 332.2 (312.0 - 354.7) | 727.8 (694.1 - 762.2) | 1378.2 (1336.1 - 1420.4) | 255.8 (237.1 - 275.7) | 441.6 (418.8 - 467.8) |

| Season | Age band | All-cause RTI hospital incidence | Lab-confirmed hMPV hospital incidence | Extrapolated hMPV hospital incidence | Lab-confirmed RSV hospital incidence | Extrapolated RSV hospital incidence | Lab-confirmed Influenza A hospital incidence | Extrapolated Influenza A hospital incidence |
|---|---|---|---|---|---|---|---|---|
| 2019/20 | <5y | 5454.6 (5363.5 - 5539.0) | 79.0 (68.2 - 89.1) | 194.3 (178.5 - 211.3) | 1004.5 (965.9 - 1044.2) | 1517.8 (1467.9 - 1564.0) | 187.8 (170.0 - 204.7) | 269.4 (248.3 - 288.8) |
| 2020/21 | <5y | 2341.4 (2284.0 - 2403.2) | 9.2 (6.0 - 13.1) | 19.2 (13.9 - 24.7) | 3.6 (1.6 - 6.0) | 6.6 (3.6 - 10.0) | 2.4 (0.8 - 4.4) | * |
| 2021/22 | <5y | 6881.0 (6784.1 - 6980.7) | 225.7 (205.9 - 244.2) | 537.0 (507.0 - 566.0) | 976.5 (936.1 - 1012.8) | 1626.3 (1576.3 - 1672.0) | 47.6 (40.0 - 56.9) | 66.5 (56.5 - 76.7) |
| 2022/23 | <5y | 4294.9 (4216.5 - 4371.0) | 56.6 (47.7 - 66.3) | 141.2 (127.8 - 155.7) | 882.4 (844.8 - 918.4) | 915.4 (880.4 - 953.6) | 180.8 (163.0 - 197.4) | 199.7 (182.8 - 217.2) |

### Supplementary Table S5: Laboratory confirmed virus proportion positive in RTI admissions in Lothian by age bands and month

| Virus | Age band | Virus proportion positive in RTI episodes | Month and year of hospital admission |
|---|---|---|---|
| hMPV | 0-5m | 0.011 | November 2017 |
| hMPV | 0-5m | 0.030 | January 2018 |
| hMPV | 0-5m | 0.054 | February 2018 |
| hMPV | 0-5m | 0.027 | March 2018 |
| hMPV | 0-5m | 0.148 | April 2018 |
| hMPV | 0-5m | 0.043 | May 2018 |
| hMPV | 0-5m | 0.059 | August 2018 |
| hMPV | 0-5m | 0.032 | October 2018 |
| hMPV | 0-5m | 0.037 | November 2018 |
| hMPV | 0-5m | 0.052 | December 2018 |
| hMPV | 0-5m | 0.112 | January 2019 |
| hMPV | 0-5m | 0.057 | February 2019 |
| hMPV | 0-5m | 0.061 | March 2019 |
| hMPV | 0-5m | 0.059 | April 2019 |
| hMPV | 0-5m | 0.030 | May 2019 |
| hMPV | 0-5m | 0.071 | June 2019 |
| hMPV | 0-5m | 0.143 | August 2019 |
| hMPV | 0-5m | 0.028 | September 2019 |
| hMPV | 0-5m | 0.021 | October 2019 |
| hMPV | 0-5m | 0.006 | November 2019 |
| hMPV | 0-5m | 0.049 | December 2019 |
| hMPV | 0-5m | 0.100 | January 2020 |
| hMPV | 0-5m | 0.083 | February 2020 |
| hMPV | 0-5m | 0.032 | March 2020 |
| hMPV | 0-5m | 0.048 | July 2021 |
| hMPV | 0-5m | 0.020 | September 2021 |
| hMPV | 0-5m | 0.030 | October 2021 |
| hMPV | 0-5m | 0.210 | November 2021 |
| hMPV | 0-5m | 0.317 | December 2021 |
| hMPV | 0-5m | 0.073 | January 2022 |
| hMPV | 0-5m | 0.019 | April 2022 |
| hMPV | 0-5m | 0.009 | October 2022 |
| hMPV | 0-5m | 0.050 | December 2022 |
| hMPV | 6m-1y | 0.053 | July 2017 |
| hMPV | 6m-1y | 0.013 | September 2017 |
| hMPV | 6m-1y | 0.065 | November 2017 |
| hMPV | 6m-1y | 0.054 | December 2017 |

| | | | |
|---|---|---|---|
| hMPV | 6m-1y | 0.026 | January 2018 |
| hMPV | 6m-1y | 0.089 | February 2018 |
| hMPV | 6m-1y | 0.044 | March 2018 |
| hMPV | 6m-1y | 0.056 | April 2018 |
| hMPV | 6m-1y | 0.109 | May 2018 |
| hMPV | 6m-1y | 0.037 | June 2018 |
| hMPV | 6m-1y | 0.095 | July 2018 |
| hMPV | 6m-1y | 0.032 | November 2018 |
| hMPV | 6m-1y | 0.068 | December 2018 |
| hMPV | 6m-1y | 0.127 | January 2019 |
| hMPV | 6m-1y | 0.081 | February 2019 |
| hMPV | 6m-1y | 0.115 | March 2019 |
| hMPV | 6m-1y | 0.029 | April 2019 |
| hMPV | 6m-1y | 0.049 | May 2019 |
| hMPV | 6m-1y | 0.036 | June 2019 |
| hMPV | 6m-1y | 0.014 | September 2019 |
| hMPV | 6m-1y | 0.033 | October 2019 |
| hMPV | 6m-1y | 0.031 | November 2019 |
| hMPV | 6m-1y | 0.030 | December 2019 |
| hMPV | 6m-1y | 0.118 | January 2020 |
| hMPV | 6m-1y | 0.048 | February 2020 |
| hMPV | 6m-1y | 0.032 | March 2020 |
| hMPV | 6m-1y | 0.091 | August 2020 |
| hMPV | 6m-1y | 0.011 | August 2021 |
| hMPV | 6m-1y | 0.019 | October 2021 |
| hMPV | 6m-1y | 0.266 | November 2021 |
| hMPV | 6m-1y | 0.342 | December 2021 |
| hMPV | 6m-1y | 0.097 | January 2022 |
| hMPV | 6m-1y | 0.034 | February 2022 |
| hMPV | 6m-1y | 0.008 | October 2022 |
| hMPV | 6m-1y | 0.026 | November 2022 |
| hMPV | 6m-1y | 0.197 | December 2022 |
| hMPV | 6m-1y | 0.062 | January 2023 |
| hMPV | 6m-1y | 0.250 | February 2023 |
| hMPV | 6m-1y | 0.022 | March 2023 |
| hMPV | 6m-1y | 0.056 | April 2023 |
| hMPV | 6m-1y | 0.059 | May 2023 |
| hMPV | 2-4y | 0.133 | July 2017 |
| hMPV | 2-4y | 0.020 | November 2017 |
| hMPV | 2-4y | 0.081 | December 2017 |
| hMPV | 2-4y | 0.118 | January 2018 |
| hMPV | 2-4y | 0.077 | February 2018 |
| hMPV | 2-4y | 0.030 | March 2018 |
| hMPV | 2-4y | 0.056 | April 2018 |

| | | | |
|---|---|---|---|
| hMPV | 2-4y | 0.091 | May 2018 |
| hMPV | 2-4y | 0.053 | June 2018 |
| hMPV | 2-4y | 0.133 | October 2018 |
| hMPV | 2-4y | 0.038 | November 2018 |
| hMPV | 2-4y | 0.066 | December 2018 |
| hMPV | 2-4y | 0.082 | January 2019 |
| hMPV | 2-4y | 0.114 | March 2019 |
| hMPV | 2-4y | 0.182 | April 2019 |
| hMPV | 2-4y | 0.133 | May 2019 |
| hMPV | 2-4y | 0.143 | June 2019 |
| hMPV | 2-4y | 0.026 | July 2019 |
| hMPV | 2-4y | 0.031 | August 2019 |
| hMPV | 2-4y | 0.057 | September 2019 |
| hMPV | 2-4y | 0.026 | October 2019 |
| hMPV | 2-4y | 0.016 | November 2019 |
| hMPV | 2-4y | 0.011 | December 2019 |
| hMPV | 2-4y | 0.056 | January 2020 |
| hMPV | 2-4y | 0.133 | February 2020 |
| hMPV | 2-4y | 0.088 | March 2020 |
| hMPV | 2-4y | 0.111 | April 2020 |
| hMPV | 2-4y | 0.023 | June 2021 |
| hMPV | 2-4y | 0.036 | July 2021 |
| hMPV | 2-4y | 0.022 | September 2021 |
| hMPV | 2-4y | 0.089 | October 2021 |
| hMPV | 2-4y | 0.200 | November 2021 |
| hMPV | 2-4y | 0.325 | December 2021 |
| hMPV | 2-4y | 0.156 | January 2022 |
| hMPV | 2-4y | 0.040 | February 2022 |
| hMPV | 2-4y | 0.043 | March 2022 |
| hMPV | 2-4y | 0.041 | September 2022 |
| hMPV | 2-4y | 0.016 | October 2022 |
| hMPV | 2-4y | 0.108 | November 2022 |
| hMPV | 2-4y | 0.044 | December 2022 |
| hMPV | 2-4y | 0.136 | January 2023 |
| hMPV | 2-4y | 0.130 | February 2023 |
| hMPV | 2-4y | 0.050 | March 2023 |
| RSV | 0-5m | 0.053 | July 2017 |
| RSV | 0-5m | 0.133 | August 2017 |
| RSV | 0-5m | 0.115 | September 2017 |
| RSV | 0-5m | 0.314 | October 2017 |
| RSV | 0-5m | 0.534 | November 2017 |
| RSV | 0-5m | 0.542 | December 2017 |
| RSV | 0-5m | 0.197 | January 2018 |
| RSV | 0-5m | 0.081 | February 2018 |

| | | | |
|---|---|---|---|
| RSV | 0-5m | 0.081 | March 2018 |
| RSV | 0-5m | 0.111 | April 2018 |
| RSV | 0-5m | 0.087 | May 2018 |
| RSV | 0-5m | 0.300 | June 2018 |
| RSV | 0-5m | 0.059 | August 2018 |
| RSV | 0-5m | 0.219 | September 2018 |
| RSV | 0-5m | 0.419 | October 2018 |
| RSV | 0-5m | 0.670 | November 2018 |
| RSV | 0-5m | 0.607 | December 2018 |
| RSV | 0-5m | 0.325 | January 2019 |
| RSV | 0-5m | 0.132 | February 2019 |
| RSV | 0-5m | 0.041 | March 2019 |
| RSV | 0-5m | 0.029 | April 2019 |
| RSV | 0-5m | 0.059 | July 2019 |
| RSV | 0-5m | 0.190 | August 2019 |
| RSV | 0-5m | 0.250 | September 2019 |
| RSV | 0-5m | 0.562 | October 2019 |
| RSV | 0-5m | 0.632 | November 2019 |
| RSV | 0-5m | 0.504 | December 2019 |
| RSV | 0-5m | 0.400 | January 2020 |
| RSV | 0-5m | 0.125 | February 2020 |
| RSV | 0-5m | 0.143 | July 2021 |
| RSV | 0-5m | 0.641 | August 2021 |
| RSV | 0-5m | 0.741 | September 2021 |
| RSV | 0-5m | 0.570 | October 2021 |
| RSV | 0-5m | 0.129 | November 2021 |
| RSV | 0-5m | 0.098 | December 2021 |
| RSV | 0-5m | 0.055 | January 2022 |
| RSV | 0-5m | 0.068 | March 2022 |
| RSV | 0-5m | 0.074 | April 2022 |
| RSV | 0-5m | 0.133 | May 2022 |
| RSV | 0-5m | 0.329 | June 2022 |
| RSV | 0-5m | 0.115 | July 2022 |
| RSV | 0-5m | 0.169 | August 2022 |
| RSV | 0-5m | 0.444 | September 2022 |
| RSV | 0-5m | 0.440 | October 2022 |
| RSV | 0-5m | 0.477 | November 2022 |
| RSV | 0-5m | 0.250 | December 2022 |
| RSV | 0-5m | 0.375 | January 2023 |
| RSV | 6m-1y | 0.026 | July 2017 |
| RSV | 6m-1y | 0.065 | August 2017 |
| RSV | 6m-1y | 0.080 | September 2017 |
| RSV | 6m-1y | 0.228 | October 2017 |
| RSV | 6m-1y | 0.336 | November 2017 |

| | | | |
|---|---|---|---|
| RSV | 6m-1y | 0.369 | December 2017 |
| RSV | 6m-1y | 0.077 | January 2018 |
| RSV | 6m-1y | 0.044 | February 2018 |
| RSV | 6m-1y | 0.015 | March 2018 |
| RSV | 6m-1y | 0.056 | April 2018 |
| RSV | 6m-1y | 0.036 | May 2018 |
| RSV | 6m-1y | 0.037 | June 2018 |
| RSV | 6m-1y | 0.143 | July 2018 |
| RSV | 6m-1y | 0.075 | August 2018 |
| RSV | 6m-1y | 0.089 | September 2018 |
| RSV | 6m-1y | 0.459 | October 2018 |
| RSV | 6m-1y | 0.529 | November 2018 |
| RSV | 6m-1y | 0.444 | December 2018 |
| RSV | 6m-1y | 0.203 | January 2019 |
| RSV | 6m-1y | 0.058 | February 2019 |
| RSV | 6m-1y | 0.090 | March 2019 |
| RSV | 6m-1y | 0.055 | June 2019 |
| RSV | 6m-1y | 0.065 | July 2019 |
| RSV | 6m-1y | 0.122 | August 2019 |
| RSV | 6m-1y | 0.229 | September 2019 |
| RSV | 6m-1y | 0.496 | October 2019 |
| RSV | 6m-1y | 0.556 | November 2019 |
| RSV | 6m-1y | 0.351 | December 2019 |
| RSV | 6m-1y | 0.197 | January 2020 |
| RSV | 6m-1y | 0.048 | February 2020 |
| RSV | 6m-1y | 0.016 | March 2020 |
| RSV | 6m-1y | 0.096 | July 2021 |
| RSV | 6m-1y | 0.521 | August 2021 |
| RSV | 6m-1y | 0.559 | September 2021 |
| RSV | 6m-1y | 0.330 | October 2021 |
| RSV | 6m-1y | 0.032 | November 2021 |
| RSV | 6m-1y | 0.017 | December 2021 |
| RSV | 6m-1y | 0.014 | January 2022 |
| RSV | 6m-1y | 0.069 | February 2022 |
| RSV | 6m-1y | 0.045 | March 2022 |
| RSV | 6m-1y | 0.180 | April 2022 |
| RSV | 6m-1y | 0.203 | May 2022 |
| RSV | 6m-1y | 0.198 | June 2022 |
| RSV | 6m-1y | 0.158 | July 2022 |
| RSV | 6m-1y | 0.189 | August 2022 |
| RSV | 6m-1y | 0.202 | September 2022 |
| RSV | 6m-1y | 0.297 | October 2022 |
| RSV | 6m-1y | 0.368 | November 2022 |
| RSV | 6m-1y | 0.098 | December 2022 |

| | | | |
|---|---|---|---|
| RSV | 6m-1y | 0.094 | January 2023 |
| RSV | 6m-1y | 0.022 | March 2023 |
| RSV | 6m-1y | 0.111 | April 2023 |
| RSV | 6m-1y | 0.118 | May 2023 |
| RSV | 2-4y | 0.100 | October 2017 |
| RSV | 2-4y | 0.327 | November 2017 |
| RSV | 2-4y | 0.145 | December 2017 |
| RSV | 2-4y | 0.059 | January 2018 |
| RSV | 2-4y | 0.030 | March 2018 |
| RSV | 2-4y | 0.036 | September 2018 |
| RSV | 2-4y | 0.300 | October 2018 |
| RSV | 2-4y | 0.410 | November 2018 |
| RSV | 2-4y | 0.355 | December 2018 |
| RSV | 2-4y | 0.020 | January 2019 |
| RSV | 2-4y | 0.075 | September 2019 |
| RSV | 2-4y | 0.224 | October 2019 |
| RSV | 2-4y | 0.317 | November 2019 |
| RSV | 2-4y | 0.126 | December 2019 |
| RSV | 2-4y | 0.023 | June 2021 |
| RSV | 2-4y | 0.036 | July 2021 |
| RSV | 2-4y | 0.611 | August 2021 |
| RSV | 2-4y | 0.516 | September 2021 |
| RSV | 2-4y | 0.107 | October 2021 |
| RSV | 2-4y | 0.012 | November 2021 |
| RSV | 2-4y | 0.013 | December 2021 |
| RSV | 2-4y | 0.020 | February 2022 |
| RSV | 2-4y | 0.022 | March 2022 |
| RSV | 2-4y | 0.139 | April 2022 |
| RSV | 2-4y | 0.157 | May 2022 |
| RSV | 2-4y | 0.179 | June 2022 |
| RSV | 2-4y | 0.205 | July 2022 |
| RSV | 2-4y | 0.130 | August 2022 |
| RSV | 2-4y | 0.178 | September 2022 |
| RSV | 2-4y | 0.219 | October 2022 |
| RSV | 2-4y | 0.169 | November 2022 |
| RSV | 2-4y | 0.067 | December 2022 |
| RSV | 2-4y | 0.045 | January 2023 |
| RSV | 2-4y | 0.043 | February 2023 |
| RSV | 2-4y | 0.150 | March 2023 |
| RSV | 2-4y | 0.125 | April 2023 |
| RSV | 2-4y | 0.111 | June 2023 |
| Influenza A | 0-5m | 0.038 | September 2017 |
| Influenza A | 0-5m | 0.029 | October 2017 |
| Influenza A | 0-5m | 0.011 | November 2017 |

| | | | |
|---|---|---|---|
| Influenza A | 0-5m | 0.084 | December 2017 |
| Influenza A | 0-5m | 0.106 | January 2018 |
| Influenza A | 0-5m | 0.027 | March 2018 |
| Influenza A | 0-5m | 0.074 | April 2018 |
| Influenza A | 0-5m | 0.007 | December 2018 |
| Influenza A | 0-5m | 0.100 | January 2019 |
| Influenza A | 0-5m | 0.094 | February 2019 |
| Influenza A | 0-5m | 0.061 | March 2019 |
| Influenza A | 0-5m | 0.048 | August 2019 |
| Influenza A | 0-5m | 0.028 | September 2019 |
| Influenza A | 0-5m | 0.010 | October 2019 |
| Influenza A | 0-5m | 0.011 | November 2019 |
| Influenza A | 0-5m | 0.081 | December 2019 |
| Influenza A | 0-5m | 0.083 | January 2020 |
| Influenza A | 0-5m | 0.018 | January 2022 |
| Influenza A | 0-5m | 0.018 | October 2022 |
| Influenza A | 0-5m | 0.023 | November 2022 |
| Influenza A | 0-5m | 0.050 | December 2022 |
| Influenza A | 0-5m | 0.125 | January 2023 |
| Influenza A | 6m-1y | 0.032 | August 2017 |
| Influenza A | 6m-1y | 0.009 | November 2017 |
| Influenza A | 6m-1y | 0.077 | December 2017 |
| Influenza A | 6m-1y | 0.141 | January 2018 |
| Influenza A | 6m-1y | 0.044 | March 2018 |
| Influenza A | 6m-1y | 0.013 | November 2018 |
| Influenza A | 6m-1y | 0.060 | December 2018 |
| Influenza A | 6m-1y | 0.271 | January 2019 |
| Influenza A | 6m-1y | 0.221 | February 2019 |
| Influenza A | 6m-1y | 0.077 | March 2019 |
| Influenza A | 6m-1y | 0.031 | November 2019 |
| Influenza A | 6m-1y | 0.134 | December 2019 |
| Influenza A | 6m-1y | 0.092 | January 2020 |
| Influenza A | 6m-1y | 0.012 | February 2020 |
| Influenza A | 6m-1y | 0.016 | March 2020 |
| Influenza A | 6m-1y | 0.060 | April 2022 |
| Influenza A | 6m-1y | 0.012 | June 2022 |
| Influenza A | 6m-1y | 0.011 | August 2022 |
| Influenza A | 6m-1y | 0.008 | October 2022 |
| Influenza A | 6m-1y | 0.079 | November 2022 |
| Influenza A | 6m-1y | 0.213 | December 2022 |
| Influenza A | 6m-1y | 0.062 | January 2023 |
| Influenza A | 6m-1y | 0.062 | February 2023 |
| Influenza A | 2-4y | 0.067 | July 2017 |
| Influenza A | 2-4y | 0.020 | November 2017 |

| | | | |
|---|---|---|---|
| Influenza A | 2-4y | 0.129 | December 2017 |
| Influenza A | 2-4y | 0.147 | January 2018 |
| Influenza A | 2-4y | 0.026 | February 2018 |
| Influenza A | 2-4y | 0.030 | March 2018 |
| Influenza A | 2-4y | 0.028 | April 2018 |
| Influenza A | 2-4y | 0.045 | May 2018 |
| Influenza A | 2-4y | 0.033 | October 2018 |
| Influenza A | 2-4y | 0.051 | November 2018 |
| Influenza A | 2-4y | 0.118 | December 2018 |
| Influenza A | 2-4y | 0.367 | January 2019 |
| Influenza A | 2-4y | 0.442 | February 2019 |
| Influenza A | 2-4y | 0.114 | March 2019 |
| Influenza A | 2-4y | 0.030 | April 2019 |
| Influenza A | 2-4y | 0.036 | June 2019 |
| Influenza A | 2-4y | 0.032 | November 2019 |
| Influenza A | 2-4y | 0.253 | December 2019 |
| Influenza A | 2-4y | 0.111 | January 2020 |
| Influenza A | 2-4y | 0.033 | February 2020 |
| Influenza A | 2-4y | 0.088 | March 2020 |
| Influenza A | 2-4y | 0.018 | October 2021 |
| Influenza A | 2-4y | 0.013 | December 2021 |
| Influenza A | 2-4y | 0.020 | February 2022 |
| Influenza A | 2-4y | 0.043 | March 2022 |
| Influenza A | 2-4y | 0.111 | April 2022 |
| Influenza A | 2-4y | 0.020 | May 2022 |
| Influenza A | 2-4y | 0.043 | August 2022 |
| Influenza A | 2-4y | 0.014 | September 2022 |
| Influenza A | 2-4y | 0.047 | October 2022 |
| Influenza A | 2-4y | 0.046 | November 2022 |
| Influenza A | 2-4y | 0.356 | December 2022 |
| Influenza A | 2-4y | 0.182 | January 2023 |

#### Supplementary Table S6: Laboratory-confirmed and extrapolated hospital incidence per 100,000 persons (with 95% CI) of hMPV, RSV and Influenza A by age bands, season and health board (Lothian and other health boards) in older adults in Scotland

| Season | Age band | Region | All-cause RTI incidence | Lab confirmed hMPV incidence | Extrapolated hMPV incidence | Lab confirmed RSV incidence | Extrapolated RSV incidence | Lab confirmed Influenza A incidence | Extrapolated Influenza A incidence |
|---|---|---|---|---|---|---|---|---|---|
| 2017/18 | 0-5m | Lothian | 5923.6 (5437.5 - 6421) | 124.3 (56.5 - 214.8) | NA (NA - NA) | 1865.2 (1582.6 - 2159.2) | NA (NA - NA) | 271.3 (169.6 - 384.4) | NA (NA - NA) |
| 2017/18 | 0-5m | Other health boards | 7498.4 (7238.5 - 7734.8) | 105.3 (74.9 - 138.1) | 192.1 (154.4 - 234) | 1661.6 (1537.6 - 1781.1) | 2316.2 (2171.7 - 2462.1) | 142.8 (107.7 - 177.9) | 321.7 (271.5 - 379.1) |
| 2018/19 | 0-5m | Lothian | 7117.8 (6602.4 - 7656.3) | 386.3 (257.6 - 515.1) | NA (NA - NA) | 2481.9 (2153.8 - 2809.9) | NA (NA - NA) | 199 (117.1 - 304.4) | NA (NA - NA) |
| 2018/19 | 0-5m | Other health boards | 7893.4 (7613.2 - 8147.4) | 102.1 (71.2 - 135.3) | 398.5 (339.5 - 455.8) | 1583.4 (1471.8 - 1709.3) | 2867.9 (2718.2 - 3019.7) | 166.2 (128.2 - 201.8) | 213.6 (175.7 - 256.4) |
| 2019/20 | 0-5m | Lothian | 7633.9 (7045.7 - 8222.3) | 294.1 (171.5 - 416.6) | NA (NA - NA) | 3308.4 (2928.6 - 3713.1) | NA (NA - NA) | 245.1 (147 - 355.3) | NA (NA - NA) |
| 2019/20 | 0-5m | Other health boards | 7643.2 (7387.5 - 7889.2) | 59.6 (37.2 - 81.9) | 258.6 (211 - 302.9) | 2301.2 (2149.6 - 2450.2) | 3230.4 (3058.3 - 3413.3) | 163.8 (124.1 - 203.6) | 275.8 (225.9 - 332.6) |
| 2020/21 | 0-5m | Lothian | 2173.9 (1858.1 - 2489.7) | * | NA (NA - NA) | * | NA (NA - NA) | * | NA (NA - NA) |
| 2020/21 | 0-5m | Other health boards | 2111 (1973.1 - 2246.1) | 10.4 (2.6 - 20.8) | † | 2.6 (0 - 7.8) | † | 2.6 (0 - 10.4) | † |
| 2021/22 | 0-5m | Lothian | 10532.7 (9874.1 - 11191.3) | 633.9 (463.2 - 804.9) | NA (NA - NA) | 3718.2 (3315.9 - 4120.4) | NA (NA - NA) | 12.2 (0 - 36.6) | NA (NA - NA) |

* There were no laboratory-confirmed episodes in Lothian in the age band and season combination.

† There were no laboratory-confirmed episodes in Lothian in the age band and season combination. As a result, extrapolation was not possible.

| | | | | | | | | | |
|---|---|---|---|---|---|---|---|---|---|
| 2021/22 | 0-5m | Other health boards | 10187.8 (9897.9 - 10464.8) | 174.4 (133.4 - 218) | 673.8 (595.1 - 756.7) | 2021.1 (1882.6 - 2172.5) | 3364.4 (3183 - 3539.6) | 43.6 (23.1 - 66.7) | 54.9 (33.3 - 79.5) |
| 2022/23 | 0-5m | Lothian | 5558.3 (5045.1 - 6046.6) | 25 (0 - 62.6) | NA (NA - NA) | 1865.3 (1589.6 - 2166.1) | NA (NA - NA) | 75.1 (25 - 137.7) | NA (NA - NA) |
| 2022/23 | 0-5m | Other health boards | 5556.3 (5307.7 - 5786.7) | 31.4 (15.7 - 49.7) | 32.7 (15.7 - 52.3) | 2174.9 (2033.5 - 2324.1) | 1881.4 (1737.8 - 2015.2) | 99.5 (70.7 - 130.9) | 95.8 (65.4 - 128.2) |
| 2017/18 | 6m-1y | Lothian | 8603.7 (8021.9 - 9140.7) | 391.6 (257.3 - 514.7) | NA (NA - NA) | 1398.5 (1152.1 - 1633.5) | NA (NA - NA) | 290.9 (179 - 402.8) | NA (NA - NA) |
| 2017/18 | 6m-1y | Other health boards | 14925.9 (14592.9 - 15267.9) | 198.3 (159.6 - 239.3) | 698.5 (620 - 781.9) | 1370 (1255.9 - 1479.4) | 2404 (2258.9 - 2553.1) | 328.2 (278 - 378.4) | 491.3 (424 - 553.9) |
| 2018/19 | 6m-1y | Lothian | 10787.7 (10191.9 - 11450.7) | 606.8 (449.5 - 764.4) | NA (NA - NA) | 2595.8 (2269.9 - 2932.9) | NA (NA - NA) | 752.9 (584.3 - 943.9) | NA (NA - NA) |
| 2018/19 | 6m-1y | Other health boards | 14390.6 (14048.1 - 14723.8) | 184 (144.4 - 226) | 828.3 (747.8 - 908.6) | 1362.9 (1253.3 - 1465.4) | 3427.8 (3254.5 - 3597.1) | 512.5 (447.3 - 575.4) | 984.6 (889.9 - 1076.3) |
| 2019/20 | 6m-1y | Lothian | 9740.4 (9132.4 - 10395.2) | 339.1 (233.9 - 467.7) | NA (NA - NA) | 2841.4 (2490.6 - 3203.9) | NA (NA - NA) | 374.2 (257.2 - 514.5) | NA (NA - NA) |
| 2019/20 | 6m-1y | Other health boards | 13780.1 (13426.6 - 14107.6) | 162.6 (124.9 - 202.6) | 451 (384.1 - 518.4) | 2014.7 (1875.6 - 2153.7) | 3982.3 (3796.1 - 4166.1) | 433.6 (367.6 - 502) | 596.1 (527.8 - 669.2) |
| 2020/21 | 6m-1y | Lothian | 4641.8 (4176.4 - 5107.5) | 24.5 (0 - 61.2) | NA (NA - NA) | * | NA (NA - NA) | * | NA (NA - NA) |
| 2020/21 | 6m-1y | Other health boards | 6827.8 (6580.6 - 7060.2) | 22.2 (9.9 - 37.1) | 62.9 (39.6 - 86.5) | 9.9 (2.5 - 19.8) | † | 2.5 (0 - 7.4) | † |
| 2021/22 | 6m-1y | Lothian | 12432.9 (11729.6 - 13148.2) | 1013.2 (810.6 - 1227.8) | NA (NA - NA) | 2670.2 (2312.6 - 3015.9) | NA (NA - NA) | 47.7 (11.9 - 95.4) | NA (NA - NA) |
| 2021/22 | 6m-1y | Other health boards | 15769.6 (15394.9 - 16136.6) | 438.3 (369.5 - 502.1) | 1241.6 (1129 - 1343.1) | 1524 (1411.8 - 1641.2) | 3421.6 (3256.9 - 3616.2) | 86.6 (58.6 - 117.3) | 116.7 (84.1 - 152.9) |
| 2022/23 | 6m-1y | Lothian | 8766.3 (8149.5 - 9395) | 296.6 (189.5 - 415.2) | NA (NA - NA) | 1755.6 (1470.9 - 2028.8) | NA (NA - NA) | 320.3 (201.7 - 450.8) | NA (NA - NA) |
| 2022/23 | 6m-1y | Other health boards | 9669.1 (9367 - 9981.3) | 119.9 (84.9 - 154.8) | 390.4 (329.6 - 449.6) | 1700.6 (1578.2 - 1823) | 1899.7 (1767.9 - 2045.2) | 379.6 (319.6 - 439.5) | 410.1 (347 - 472) |
| 2017/18 | 2-4y | Lothian | 1389.8 (1265.4 - 1524.9) | 74.6 (46.2 - 106.7) | NA (NA - NA) | 110.2 (74.6 - 149.4) | NA (NA - NA) | 67.5 (39.1 - 99.5) | NA (NA - NA) |

| | | | | | | | | | |
|---|---|---|---|---|---|---|---|---|---|
| 2017/18 | 2-4y | Other health boards | 2522.1 (2441.6 - 2604) | 34.5 (24.4 - 43.8) | 141.1 (120 - 161.7) | 140.2 (121.4 - 159.6) | 217.3 (194.1 - 243.7) | 78.3 (64 - 92.7) | 114.6 (96.3 - 132.3) |
| 2018/19 | 2-4y | Lothian | 1635 (1501.1 - 1786.9) | 115.7 (79.6 - 155.6) | NA (NA - NA) | 253.2 (191.7 - 314.7) | NA (NA - NA) | 220.6 (166.4 - 278.5) | NA (NA - NA) |
| 2018/19 | 2-4y | Other health boards | 2698.1 (2608.2 - 2789.5) | 34.3 (24.8 - 44.6) | 178.6 (157.1 - 201.7) | 130.1 (112.5 - 149.1) | 355.9 (324.5 - 388.8) | 181.2 (157.1 - 203.2) | 381 (348.6 - 412.9) |
| 2019/20 | 2-4y | Lothian | 1767.2 (1604.5 - 1926.1) | 70.2 (40.7 - 103.5) | NA (NA - NA) | 195.9 (144.2 - 251.4) | NA (NA - NA) | 125.7 (84.9 - 166.4) | NA (NA - NA) |
| 2019/20 | 2-4y | Other health boards | 2472.6 (2386 - 2556.2) | 30.1 (21.1 - 40.6) | 102.5 (86.5 - 121.2) | 193.4 (170.9 - 217.5) | 285.2 (256.7 - 312.4) | 113.7 (95.6 - 131.7) | 187.1 (165.6 - 210) |
| 2020/21 | 2-4y | Lothian | 674.8 (578.9 - 782.1) | 3.8 (0 - 11.5) | NA (NA - NA) | 3.8 (0 - 11.5) | NA (NA - NA) | * | NA (NA - NA) |
| 2020/21 | 2-4y | Other health boards | 1210.3 (1150 - 1274.3) | 5.4 (1.5 - 9.3) | 12.1 (6.9 - 19.3) | 2.3 (0 - 5.4) | 8.2 (3.9 - 13.1) | 3.1 (0.8 - 6.2) | † |
| 2021/22 | 2-4y | Lothian | 2640.8 (2461 - 2840.1) | 234.4 (175.8 - 293.1) | NA (NA - NA) | 453.2 (382.7 - 535.2) | NA (NA - NA) | 39.1 (15.6 - 62.5) | NA (NA - NA) |
| 2021/22 | 2-4y | Other health boards | 3380 (3288.9 - 3473.5) | 95.8 (79.3 - 113.9) | 301.2 (271 - 331.5) | 304.8 (274.1 - 334.6) | 572.9 (532.5 - 612.7) | 40.8 (29.8 - 51.8) | 64.9 (50.3 - 79.3) |
| 2022/23 | 2-4y | Lothian | 1681.8 (1519 - 1844.6) | 77.5 (46.5 - 116.3) | NA (NA - NA) | 244.1 (186 - 306.1) | NA (NA - NA) | 112.4 (73.6 - 155) | NA (NA - NA) |
| 2022/23 | 2-4y | Other health boards | 2372.4 (2289.6 - 2456.8) | 26 (17.4 - 35.5) | 105.2 (87.5 - 123.8) | 244.5 (215.3 - 271.3) | 334.4 (303.6 - 363.6) | 153.8 (133.3 - 174.3) | 182.2 (159.3 - 205.1) |
| 2017/18 | <5y | Lothian | 3667.5 (3491 - 3839.5) | 145.9 (111.1 - 185.1) | NA (NA - NA) | 699.1 (622.8 - 773.1) | NA (NA - NA) | 150.3 (115.4 - 187.3) | NA (NA - NA) |
| 2017/18 | <5y | Other health boards | 5874.7 (5776.8 - 5976.6) | 79.7 (67.8 - 91.3) | 259.1 (238.3 - 280.9) | 667.2 (635.3 - 702.6) | 1039.5 (998.1 - 1081.8) | 139.1 (124 - 155.1) | 227 (207.3 - 246.8) |
| 2018/19 | <5y | Lothian | 4480.2 (4289.4 - 4664.4) | 263.9 (217.4 - 310.5) | NA (NA - NA) | 1137.8 (1040.2 - 1235.4) | NA (NA - NA) | 321.6 (272.8 - 381.5) | NA (NA - NA) |
| 2018/19 | <5y | Other health boards | 5946.3 (5850.7 - 6041) | 76.2 (64.9 - 87.4) | 346 (322.2 - 369.6) | 644.5 (610.7 - 678.7) | 1427.1 (1376.4 - 1474.2) | 242.5 (221.3 - 262.3) | 466 (439.9 - 492.6) |
| 2019/20 | <5y | Lothian | 4419.4 (4234.2 - 4618.2) | 164.5 (130.2 - 205.7) | NA (NA - NA) | 1293.4 (1186 - 1407.6) | NA (NA - NA) | 196.5 (153.1 - 239.9) | NA (NA - NA) |

| | | | | | | | | | |
|---|---|---|---|---|---|---|---|---|---|
| 2019/20 | <5y | Other health boards | 5664.7 (5572.9 - 5758.4) | 61.7 (51.5 - 71.9) | 200.3 (182.3 - 218.5) | 945.8 (905 - 983.9) | 1563.3 (1509 - 1614.2) | 186 (167.9 - 206) | 284.2 (261.6 - 307.6) |
| 2020/21 | <5y | Lothian | 1727.8 (1614.7 - 1852.5) | 8.8 (0 - 20.4) | NA (NA - NA) | 3.8 (0 - 15.3) | NA (NA - NA) | * | NA (NA - NA) |
| 2020/21 | <5y | Other health boards | 2466.5 (2403.1 - 2530.8) | 9.6 (5.8 - 13.9) | 21.6 (15.8 - 28.3) | 3.8 (1.4 - 6.7) | 7.5 (4.3 - 11) | 2.9 (1 - 5.3) | † |
| 2021/22 | <5y | Lothian | 6122.3 (5892.3 - 6357) | 466.9 (405.3 - 526.3) | NA (NA - NA) | 1528.8 (1407.9 - 1644.9) | NA (NA - NA) | 35.6 (19 - 54.5) | NA (NA - NA) |
| 2021/22 | <5y | Other health boards | 7036.7 (6918.9 - 7142.9) | 176.1 (158.1 - 193.6) | 551.4 (519.6 - 584.8) | 863.1 (827.1 - 904.5) | 1646.3 (1595.3 - 1701.4) | 50.1 (40.9 - 60.3) | 72.9 (60.8 - 84.2) |
| 2022/23 | <5y | Lothian | 3829.6 (3640 - 4028.5) | 111.3 (80.5 - 142.1) | NA (NA - NA) | 852.6 (767.3 - 940.2) | NA (NA - NA) | 146.8 (111.3 - 187.1) | NA (NA - NA) |
| 2022/23 | <5y | Other health boards | 4390.8 (4309.3 - 4484.9) | 45.4 (36.6 - 54.1) | 147.4 (131.7 - 162.4) | 888.6 (850 - 926.6) | 928.4 (886.6 - 968.6) | 187.8 (169.2 - 206.8) | 210.6 (190.2 - 230.7) |

#### Supplementary Table S7: Findings of sensitivity analysis conducted by limiting to episodes with specimens collected for viral testing between 3 days before and 3 days after the date of admission in older adults in Lothian by age bands and seasons

| Season | Age band | Lab-confirmed hMPV hospital incidence (sensitivity analysis) | Lab-confirmed hMPV hospital incidence (main analysis) | Lab-confirmed RSV hospital incidence (sensitivity analysis) | Lab-confirmed RSV hospital incidence (main analysis) | Lab-confirmed Influenza A hospital incidence (sensitivity analysis) | Lab-confirmed Influenza A hospital incidence (main analysis) |
|---|---|---|---|---|---|---|---|
| 2017/18 | 0-5m | 124.3 (56.5 - 203.5) | 124.3 (56.5 - 214.8) | 1831.3 (1571.3 - 2102.9) | 1865.2 (1582.6 - 2159.2) | 237.4 (135.7 - 339.1) | 271.3 (169.6 - 384.4) |
| 2018/19 | 0-5m | 386.3 (257.6 - 526.8) | 386.3 (257.6 - 515.1) | 2435 (2107.2 - 2739.4) | 2481.9 (2153.8 - 2809.9) | 199 (105.4 - 304.4) | 199 (117.1 - 304.4) |
| 2019/20 | 0-5m | 281.8 (171.5 - 392.1) | 294.1 (171.5 - 416.6) | 3210.4 (2830.2 - 3614.8) | 3308.4 (2928.6 - 3713.1) | 245.1 (147 - 355.3) | 245.1 (147 - 355.3) |
| 2020/21 | 0-5m | * | * | * | * | * | * |
| 2021/22 | 0-5m | 633.9 (463.2 - 816.8) | 633.9 (463.2 - 804.9) | 3681.6 (3279.3 - 4071.7) | 3718.2 (3315.9 - 4120.4) | 12.2 (0 - 36.6) | 12.2 (0 - 36.6) |
| 2022/23 | 0-5m | 25 (0 - 62.6) | 25 (0 - 62.6) | 1852.8 (1577.1 - 2153.2) | 1865.3 (1589.6 - 2166.1) | 75.1 (25 - 137.7) | 75.1 (25 - 137.7) |
| 2017/18 | 6m-1y | 358 (223.8 - 492.3) | 391.6 (257.3 - 514.7) | 1387.3 (1163.6 - 1633.8) | 1398.5 (1152.1 - 1633.5) | 279.7 (179 - 391.6) | 290.9 (179 - 402.8) |
| 2018/19 | 6m-1y | 584.3 (427 - 752.9) | 606.8 (449.5 - 764.4) | 2573.3 (2269.9 - 2910.7) | 2595.8 (2269.9 - 2932.9) | 707.9 (528.1 - 887.7) | 752.9 (584.3 - 943.9) |
| 2019/20 | 6m-1y | 339.1 (233.9 - 467.7) | 339.1 (233.9 - 467.7) | 2806.4 (2455.6 - 3157.4) | 2841.4 (2490.6 - 3203.9) | 350.8 (222.2 - 479.4) | 374.2 (257.2 - 514.5) |

* There were no lab-confirmed cases in Lothian in the age band and season combination.

| | | | | | | | |
|---|---|---|---|---|---|---|---|
| 2020/21 | 6m-1y | 24.5 (0 - 61.2) | 24.5 (0 - 61.2) | * | * | * | * |
| 2021/22 | 6m-1y | 1013.2 (798.7 - 1227.8) | 1013.2 (810.6 - 1227.8) | 2622.5 (2288.7 - 2980.1) | 2670.2 (2312.6 - 3015.9) | 47.7 (11.9 - 95.4) | 47.7 (11.9 - 95.4) |
| 2022/23 | 6m-1y | 284.7 (177.9 - 403.3) | 296.6 (189.5 - 415.2) | 1743.8 (1482.8 - 2028.5) | 1755.6 (1470.9 - 2028.8) | 320.3 (201.7 - 450.8) | 320.3 (201.7 - 450.8) |
| 2017/18 | 2-4y | 71.1 (42.7 - 103.1) | 74.6 (46.2 - 106.7) | 106.6 (71.1 - 149.3) | 110.2 (74.6 - 149.4) | 67.5 (39.1 - 99.5) | 67.5 (39.1 - 99.5) |
| 2018/19 | 2-4y | 104.9 (68.7 - 148.3) | 115.7 (79.6 - 155.6) | 246 (184.5 - 307.5) | 253.2 (191.7 - 314.7) | 220.6 (166.4 - 278.5) | 220.6 (166.4 - 278.5) |
| 2019/20 | 2-4y | 70.2 (40.7 - 103.5) | 70.2 (40.7 - 103.5) | 192.2 (140.5 - 247.7) | 195.9 (144.2 - 251.4) | 125.7 (84.9 - 166.4) | 125.7 (84.9 - 166.4) |
| 2020/21 | 2-4y | 3.8 (0 - 11.5) | 3.8 (0 - 11.5) | 3.8 (0 - 15.3) | 3.8 (0 - 11.5) | * | * |
| 2021/22 | 2-4y | 230.5 (171.9 - 289.1) | 234.4 (175.8 - 293.1) | 445.3 (371.1 - 531.3) | 453.2 (382.7 - 535.2) | 39.1 (15.6 - 62.5) | 39.1 (15.6 - 62.5) |
| 2022/23 | 2-4y | 73.6 (42.6 - 108.5) | 77.5 (46.5 - 116.3) | 240.3 (186 - 298.4) | 244.1 (186 - 306.1) | 112.4 (73.6 - 155) | 112.4 (73.6 - 155) |
| 2017/18 | <5y | 137.2 (102.4 - 172) | 145.9 (111.1 - 185.1) | 688.2 (616.3 - 766.6) | 699.1 (622.8 - 773.1) | 104.6 (65.9 - 143.4) | 150.3 (115.4 - 187.3) |
| 2018/19 | <5y | 252.8 (208.5 - 301.6) | 263.9 (217.4 - 310.5) | 1120.1 (1024.6 - 1222.1) | 1137.8 (1040.2 - 1235.4) | 141.6 (106.7 - 176.4) | 321.6 (272.8 - 381.5) |
| 2019/20 | <5y | 162.2 (128 - 201.1) | 164.5 (130.2 - 205.7) | 1265.9 (1160.8 - 1368.8) | 1293.4 (1186 - 1407.6) | 310.5 (259.5 - 361.5) | 196.5 (153.1 - 239.9) |
| 2020/21 | <5y | 8.8 (0 - 20.4) | 8.8 (0 - 20.4) | 3.8 (0 - 11.5) | 3.8 (0 - 15.3) | 189.7 (150.8 - 233.1) | * |
| 2021/22 | <5y | 464.6 (402.9 - 528.6) | 466.9 (405.3 - 526.3) | 1507.5 (1396 - 1623.7) | 1528.8 (1407.9 - 1644.9) | 35.6 (19 - 54.5) | 35.6 (19 - 54.5) |
| 2022/23 | <5y | 106.6 (78.2 - 137.4) | 111.3 (80.5 - 142.1) | 845.5 (757.9 - 930.8) | 852.6 (767.3 - 940.2) | 142.1 (106.6 - 180) | 146.8 (111.3 - 187.1) |

### Supplementary Table S8: Characteristics of all-cause RTI, hMPV, RSV, and Influenza A hospital episodes in older adults in Scotland (2017-2023)

| | | **All-cause RTI (n = 77,847)** | **hMPV (n = 1,462)** | **RSV (n = 10,985)** | **Influenza A (n = 2,124)** |
|---|---|---|---|---|---|
| **Age** | 0-5m | 19,757 (25.38%) | 318 (21.75%) | 5,025 (45.74%) | 321 (15.11%) |
| | 6m-1y | 36,118 (46.40%) | 694 (47.47%) | 4,295 (39.10%) | 891 (41.95%) |
| | 2-4y | 21,972 (28.22) | 450 (30.78%) | 1,665 (15.16%) | 912 (42.94%) |
| **Sex** | Female | 32,670 (41.97%) | 615 (42.07%) | 4,867 (44.31%) | 949 (44.68%) |
| | Male | 45,177 (58.03%) | 847 (57.93%) | 6,118 (55.69%) | 1,175 (55.32%) |
| **Ethnicity** | White | 199,926 (86.36%) | 1,085 (74.21%) | 7,541 (68.65%) | 1,524 (71.75%) |
| | Non-White | 29,593 (12.78%) | 350 (23.94%) | 2,620 (23.85%) | 471 (22.18%) |
| | Data unavailable | 1,989 (0.86%) | 27 (1.85%) | 824 (7.50%) | 129 (6.07%) |
| **SIMD** † | 1 | 21,853 (28.07%) | 319 (21.82%) | 2.936 (26.73%) | 621 (29.24%) |
| | 2 | 16,819 (21.61%) | 321 (21.96%) | 2,335 (21.26%) | 478 (22.50%) |
| | 3 | 13,685 (17.58%) | 288 (19.70%) | 1.986 (18.08%) | 364 (17.14%) |
| | 4 | 13,923 (17.89%) | 275 (18.81%) | 2,036 (18.53%) | 354 (16.67%) |
| | 5 | 111,41 (14.31%) | 255 (17.44%) | 1,641 (14.94%) | 296 (13.94%) |
| | Data unavailable | 426 (0.55%) | 4 (0.27%) | 51 (0.46%) | 11 (0.52%) |

† SIMD ranks data zones in Scotland in order of deprivation. The data are presented as SIMD quintiles with 1 indicating the most deprived area and higher quintiles progressively representing the less deprived areas

### Supplementary Table S9: Number and proportion of laboratory-confirmed hMPV, RSV and Influenza A RTI episodes requiring long hospital length of stay (LOS) in children aged <5 years in Scotland by age bands during six annual seasons (2017-2023)

| **Age bands** | **Region** | **Laboratory-confirmed hMPV RTI hospital episodes requiring long LOS** | **Laboratory-confirmed RSV RTI hospital episodes requiring long LOS** | **Laboratory-confirmed Influenza A RTI hospital episodes requiring long LOS** |
|---|---|---|---|---|
| **0-5 months** | Lothian | 16 (13.1%) | 133 (12.1%) | 5 (7.4%) |
| | Other health boards | 20 (10.2%) | 344 (8.8%) | 7 (2.8%) |
| | Overall (Scotland) | 36 (11.3%) | 477 (9.5%) | 12 (3.7%) |
| **6 months – 1 year** | Lothian | 35 (15.2%) | 83 (8.5%) | 12 (7.7%) |
| | Other health boards | 35 (7.5%) | 183 (5.5%) | 31 (4.2%) |
| | Overall (Scotland) | 70 (10.1%) | 266 (6.2%) | 43 (4.8%) |
| **2 – 4 years** | Lothian | 19 (15.0%) | 30 (9.0%) | 6 (3.9%) |
| | Other health boards | 19 (6.4%) | 93 (7.0%) | 39 (5.1%) |
| | Overall (Scotland) | 38 (8.4%) | 123 (7.4%) | 45 (4.9%) |
| **Overall (<5 years)** | Lothian | 70 (13.9%) | 246 (10.0%) | 23 (6.1%) |
| | Other health boards | 74 (7.7%) | 620 (7.2%) | 77 (4.4%) |
| | Overall (Scotland) | 98 (9.8%) | 866 (7.9%) | 100 (4.7%) |

### Supplementary Table S10: ICU admission rate and in-hospital and post-discharge CFR associated with laboratory-confirmed hMPV, RSV, and Influenza A hospital admissions

| | Laboratory-confirmed hMPV RTI hospital episodes | Laboratory-confirmed RSV RTI hospital episodes | Laboratory-confirmed Influenza A hospital episodes |
|---|---|---|---|
| **ICU admission rate** | 0.34 (0.07 – 0.68) | 0.12 (0.06 – 0.18) | 0.05 (0 – 0.19) |
| **In-hospital CFR** | 0.14 (0 – 0.34) | 0.01 (0 – 0.03) | 0 (0 – 0) |
| **90 days post-discharge CFR** | 0.34 (0.07 – 0.68) | 0.05 (0.02 – 0.10) | 0.05 (0 – 0.14) |
